# Scoring the NIH Stroke Scale from bedside exam transcripts: a synthetic benchmark of a decision model and a large language model

**DOI:** 10.64898/2026.10.23.26366376

**Authors:** Noor Abdulla, Waleed Abood

**Affiliations:** Department of Neurology, Wayne State University School of Medicine / Detroit Medical Center, Detroit, Michigan, USA

## Abstract

**Background:** Clinicians score the NIH Stroke Scale (NIHSS) at the bedside, often under time pressure, and ambient AI tools that listen to clinical encounters are now entering practice. It is not known whether software can score the NIHSS from the exam conversation itself, rather than extract a score that has already been written in a note.

**Methods:** We created 112 synthetic code-stroke cases. One hundred were allocated in proportion to real code-stroke presentations, and 12 stress cases tested special scoring rules. For each case, a large language model (Claude Opus 5.5) wrote a reference score form (the answer key) and a speech-to-text-style exam transcript, with main-set transcripts split evenly between a narrated and a telegraphic examiner style. A second model (Claude Fable 5.1), blind to the key, scored every transcript and agreed with the key on 99.2% of items. Each transcript was then scored by Jev, a decision model, and by gpt-6-luna, a general large language model (LLM). The primary outcome was item-level accuracy against the key.

**Results:** The LLM scored 99.0% of items correctly (95% CI 98.4–99.4) and Jev 94.4% (93.2–95.4; McNemar p < 0.001), with exact totals in 101 and 63 of 112 cases. Jev was faster (median 0.15 vs 5.78 s per exam) and cheaper (USD 0.23 vs 0.67 per 1,000 exams). With telegraphic examiners, accuracy fell from 98.0% to 90.9% for Jev and from 100% to 97.7% for the LLM. In a post-hoc analysis that kept only the examiner’s words, accuracy stayed high when the examiner narrated (Jev 96.9%, LLM 98.8%) but dropped to about 80% when the examiner used shorthand. Most of Jev’s errors fell on four items (language, extinction and neglect, LOC questions and visual fields), and on each it tended to repeat the same mistake, for example scoring slurred speech as aphasia. Its confidence flagged most of these errors. Re-checking the 14.8% of items with confidence below 0.70 would have caught 86% of them, and a simulated hybrid reached 98.8% accuracy.

**Conclusions:** Both engines scored most NIHSS items correctly from exam transcripts. The LLM was more accurate, and the decision model was faster and cheaper, with a confidence signal that could route uncertain items to an LLM. Because the cases and the key are synthetic, these findings need confirmation with recorded exams and human raters.

## Introduction

The NIHSS is the standard measure of stroke severity and informs acute treatment decisions [1]. Its scoring rules take formal training to apply, and even certified raters disagree. Among 8,214 raters scoring the certification videos, item-level kappa ranged from 0.15 for limb ataxia to 0.81 for level-of-consciousness commands [2], and a newer animated certification system showed a wide spread, from 0.25 to 0.90 [3].

Ambient AI scribes are being adopted to reduce documentation burden. Evidence from the emergency department is still limited, and reported problems include background noise and note accuracy [4]. Work with large language models (LLMs) on the NIHSS has so far focused on extracting scores already documented in notes, and in 312 discharge summaries, frontier LLMs reached 88.5% item accuracy [5]. To our knowledge, no study has tested scoring the NIHSS from the exam conversation itself, where each item has to be inferred from what the examiner and patient say and do.

Two kinds of engine could do this. A general LLM is flexible but relatively slow and costly per exam. A decision model answers constrained multiple-choice questions quickly and returns a confidence score for each answer. Testing either one requires exams with a known correct score. Synthetic benchmarks provide this and allow rare items and special rules to be included deliberately. They have already been used for triage scenarios [6] and doctor–patient conversations [7].

In this study, we compared a decision model (Jev) with an LLM (gpt-6-luna) for scoring the NIHSS from synthetic code-stroke exam transcripts with a known answer key. We measured item-level accuracy, agreement on the total score, speed and cost, looked at where each engine went wrong, and asked whether the decision model’s confidence could point to its own errors.

## Methods

### Study design

This was a diagnostic-accuracy study using synthetic data. We built 112 simulated code-stroke encounters with known NIHSS scores, wrote each one as a bedside exam transcript, and measured how well two scoring engines recovered the item scores. Because no patients or patient data were involved, IRB review was not required.

### Case design

We first built a literature-based table of code-stroke presentations, listing 43 presentation types with their population frequencies and typical NIHSS item profiles (69 references). Stroke mimics were left out because the study tests NIHSS scoring, not stroke diagnosis. From this table we allocated a main set of 100 cases across 31 syndrome rows in proportion to real code-stroke populations: 66 ischemic strokes (MCA/ACA cortical branch 23, lacunar 17, posterior circulation 15, large MCA territory 11), 20 TIAs, 12 intracerebral hemorrhages and 2 other hemorrhages. These ischemic groups correspond to the Oxfordshire partial anterior, lacunar, posterior and total anterior circulation categories [8]. Each case’s target NIHSS total was drawn at random from the usual range for its syndrome, with lower scores slightly more likely, as in real code-stroke populations. A fixed random seed (20261017) makes this selection reproducible.

A separate stress set of 12 cases covered rare item levels and special NIHSS rules, including bilateral blindness, intubation (dysarthria untestable), limb amputation (untestable limbs), the coma rules, locked-in syndrome, global aphasia and ataxia in two limbs. Stress cases are included in all accuracy analyses (n = 112), and figures restricted to the 100 main cases are labeled as such.

### Score forms

The score form served as the answer key for each case and was written before any transcript existed. For every case, Claude Opus 5.5 (Anthropic; claude-opus-5-5, via API) was given the syndrome, lesion side and target total. It returned the correct score for all 15 NIHSS items, a short case description (vignette) and a one-sentence reason for each abnormal item (for example, “5b = 3: right arm falls to the bed, no effort against gravity”). The prompt asked the model to follow the official NIH Stroke Scale scoring instructions, including the special rules for coma (1a = 3), aphasia, intubation and amputation or joint fusion (scored UN), limb ataxia (scored only when out of proportion to weakness) and scoring of the first attempt. All prompts and full responses were saved. A simple automated check then screened each form for internal errors, such as item scores that did not add up to the stated total, a total more than 2 points from the target, values not allowed for an item, coma-rule violations, weakness on the same side as a hemispheric lesion, aphasia with a right hemispheric lesion, or an abnormal item without a stated reason. The check flagged problems but did not change any score.

### Exam transcripts

Claude Opus 5.5 then wrote one transcript per score form (Message Batches API, temperature 1.0). Transcripts were written in a raw speech-to-text style, with timestamps, speaker labels and emergency-department background such as family members, nurses and overhead pages. They never stated a score or named an NIHSS item. Language and articulation findings came through the patient’s own speech, and slurred words were spelled phonetically (for example, “tip-tob” for “tip-top”). In 48 of the 52 cases with dysarthria, the examiner also mentioned it aloud. Examiner style was assigned at random, using the same fixed seed. In the main set, 50 transcripts used a narrated style, in which the examiner describes findings, and 50 a telegraphic style, with clipped shorthand and most findings implied. All stress cases were narrated. We fixed the encounter month to October 2026 so that the correct answer to the month question was the same in every case. The median transcript length was 689 words (range 451–986). We did not use any OpenAI model to build the cases, transcripts or answer key, because the LLM under test is an OpenAI model.

**Figure 1.**
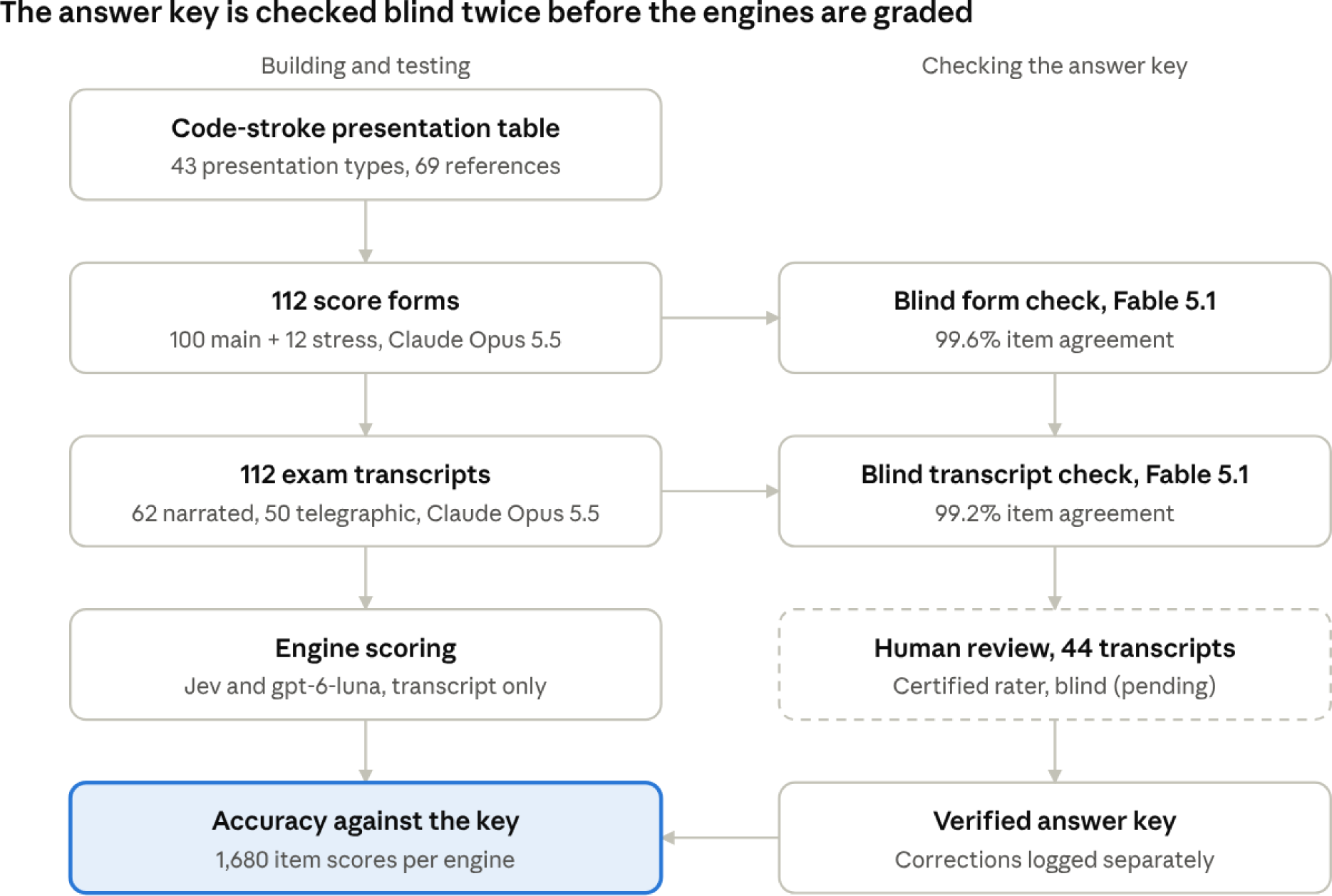
Study flow. Cases were built in the left column and the answer key was checked in the right column; dashed boxes are steps still pending. The engines saw only the transcripts and were graded against the checked key.

### Reference standard

A second model, Claude Fable 5.1 (claude-fable-5-1), checked the answer key twice without ever seeing the original scores.

1. **Score-form check.** In one call, the model rewrote each form’s vignette and reasons as plain bedside findings, with all item numbers and scores removed. In a separate call, it scored the NIHSS from those findings alone. Fable agreed with the forms on 99.6% of items (1674/1680; pooled kappa 0.99). All 15 items agreed in 107 of 112 forms, and totals never differed by more than 1 point.
2. **Transcript check.** Fable scored every transcript blind, using the transcript alone. It agreed with the key on 99.2% of items (1666/1680; kappa 0.98), and all 15 items agreed in 99 of 112 transcripts (narrated 58/62, telegraphic 41/50).
3. **Planned human review (not part of this version).** Two NIHSS-certified raters will independently score a prespecified set of 44 transcripts while blind to the key, and will rate each transcript’s realism on a 1–5 scale. The set includes all 13 transcripts with any disagreement in the transcript check, the remaining 11 stress cases, and 20 randomly chosen main-set transcripts on which the models agreed (a seeded audit). Cases are shown under random codes and mixed so that raters cannot tell which group a case comes from. A senior neurologist will resolve disagreements as an error in the form (key corrected), an unclear transcript (transcript rewritten) or a genuine judgment call (item labeled borderline). All corrections will be logged separately, and no output file will be edited by hand. Results will be reported in a revised version.

The audit sample will estimate how often agreement between the two models hides an error in the key.

### Index tests

Each transcript was scored by two engines using the same item rubric, which offered every NIHSS level, UN where the scale allows it, and a “not assessed” option:

- **Jev** (TypeSafe decision model; jev-1.13.0), which answers each item as a multiple-choice question and returns a confidence for its choice.
- **gpt-6-luna** (OpenAI), prompted with the rubric and returning structured JSON with a choice and a self-reported confidence per item.

Engine calls, rubric and settings were the same as in an earlier unpublished pilot, and each engine saw only the transcript. Latency was measured for each exam from request to response, and cost was calculated from token usage and published prices.

### Outcomes and statistics

The primary outcome was item-level accuracy, defined as the proportion of the 1,680 item scores (112 cases × 15 items) that matched the key, with Wilson 95% confidence intervals. Secondary outcomes were exact agreement on the total score, totals within 1 point, mean absolute error of the total, per-item accuracy, accuracy by examiner style and case set, latency and cost per 1,000 exams. We compared the engines on paired items with McNemar’s test. Because the 15 items within a transcript are not independent, we also compared them case by case, using the Wilcoxon signed-rank test for per-case accuracy and a case-level bootstrap (10,000 resamples of the 112 cases) for the 95% confidence interval of the accuracy difference.

In an exploratory post-hoc analysis, we simulated a hybrid system that kept Jev’s answer when its confidence reached a threshold and otherwise used the LLM’s answer for that item.

### Examiner-only analysis

After the main run, we tested how much the engines relied on speech from people other than the examiner, to approximate what a single microphone worn by the physician would capture. A script removed every labeled line (patient, nurse, family, overhead pages) from each transcript, leaving only the examiner’s words. Of 6,366 lines, 3,715 were kept, and every transcript kept at least 25 lines. Both engines scored these examiner-only transcripts with the same prompts and settings. Because TypeSafe serves only its current Jev release, which had changed from jev-1.13.0 to jev-1.14.0, we re-scored the full transcripts with jev-1.14.0 for this comparison. On the full transcripts, the two Jev versions had almost identical item accuracy (94.4% and 94.5%). This analysis was not planned in advance.

## Results

### Case set

All 112 score forms and transcripts were generated without failures. The median NIHSS in the main set was 4 (range 0–33).

**Table 1.**
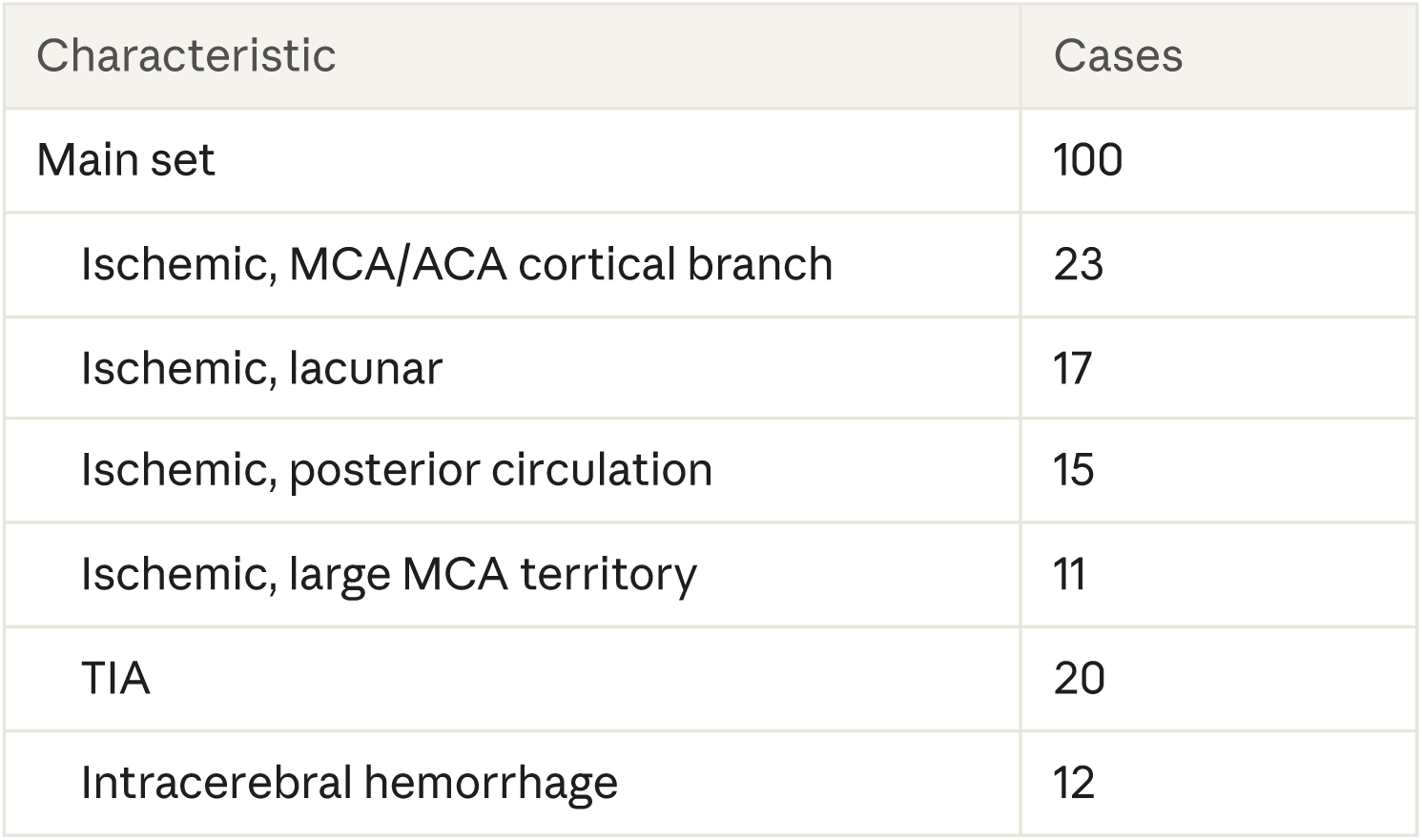
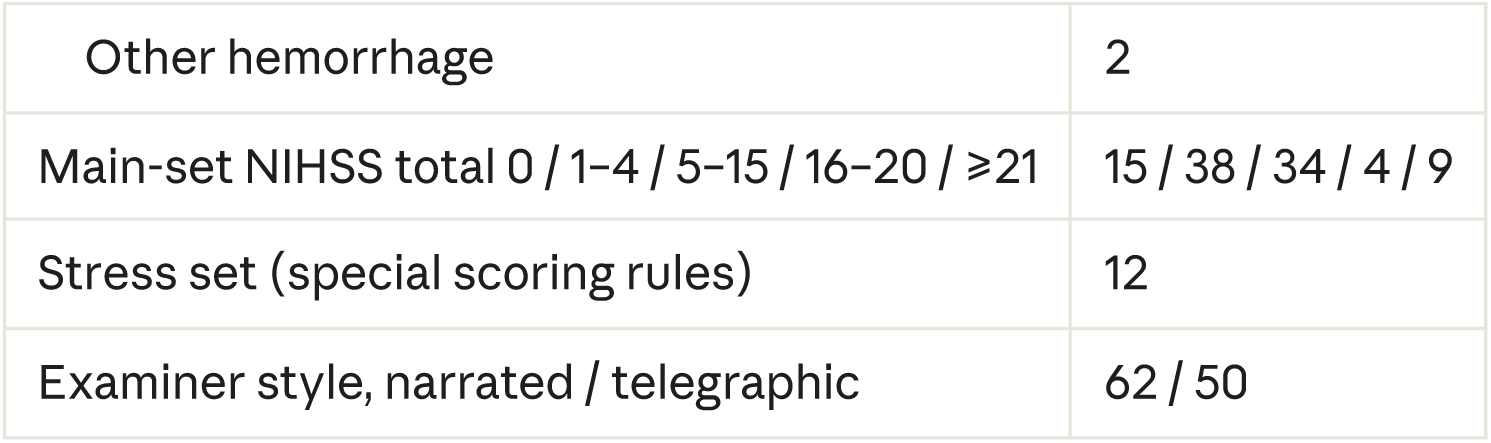
Case set.

### Transcript characteristics

Every transcript contained a full bedside exam along with the background activity of a real code stroke. All 112 included a nurse and a family member or bystander, and 109 included an overhead page, pager or alarm. No transcript stated a score or named an NIHSS item. The automated check for score leakage flagged a single line, which on review was ordinary examiner speech. Telegraphic transcripts were shorter (median 556 vs 805 words) but had as many patient turns, so the findings rested more on what the patient said and did than on the examiner’s description.

**Table 2.** Transcript characteristics by examiner style, median (range).

| Measure | Narrated (n = 62) | Telegraphic (n = 50) | All (n = 112) |
| --- | --- | --- | --- |
| Words | 805 (613–986) | 556 (451–670) | 689 (451–986) |
| Lines (utterances) | 55 (42–67) | 59 (47–73) | 56 (42–73) |
| Examiner lines | 33 (25–42) | 33 (25–45) | 33 (25–45) |
| Patient lines | 15 (0–22) | 19 (0–29) | 17 (0–29) |
| Distinct speakers | 5 (3–6) | 5 (4–6) | 5 (3–6) |
| Simulated exam length, min:s | 3:49 (3:02–4:56) | 3:18 (2:33–4:01) | 3:36 (2:33–4:56) |
| With a page, pager or alarm | 62 | 47 | 109 |

Six transcripts had no patient lines because the patient could not speak (global aphasia, stupor, intubation or locked-in syndrome). Simulated length is the time stamp of the last line.

**Figure 2.**
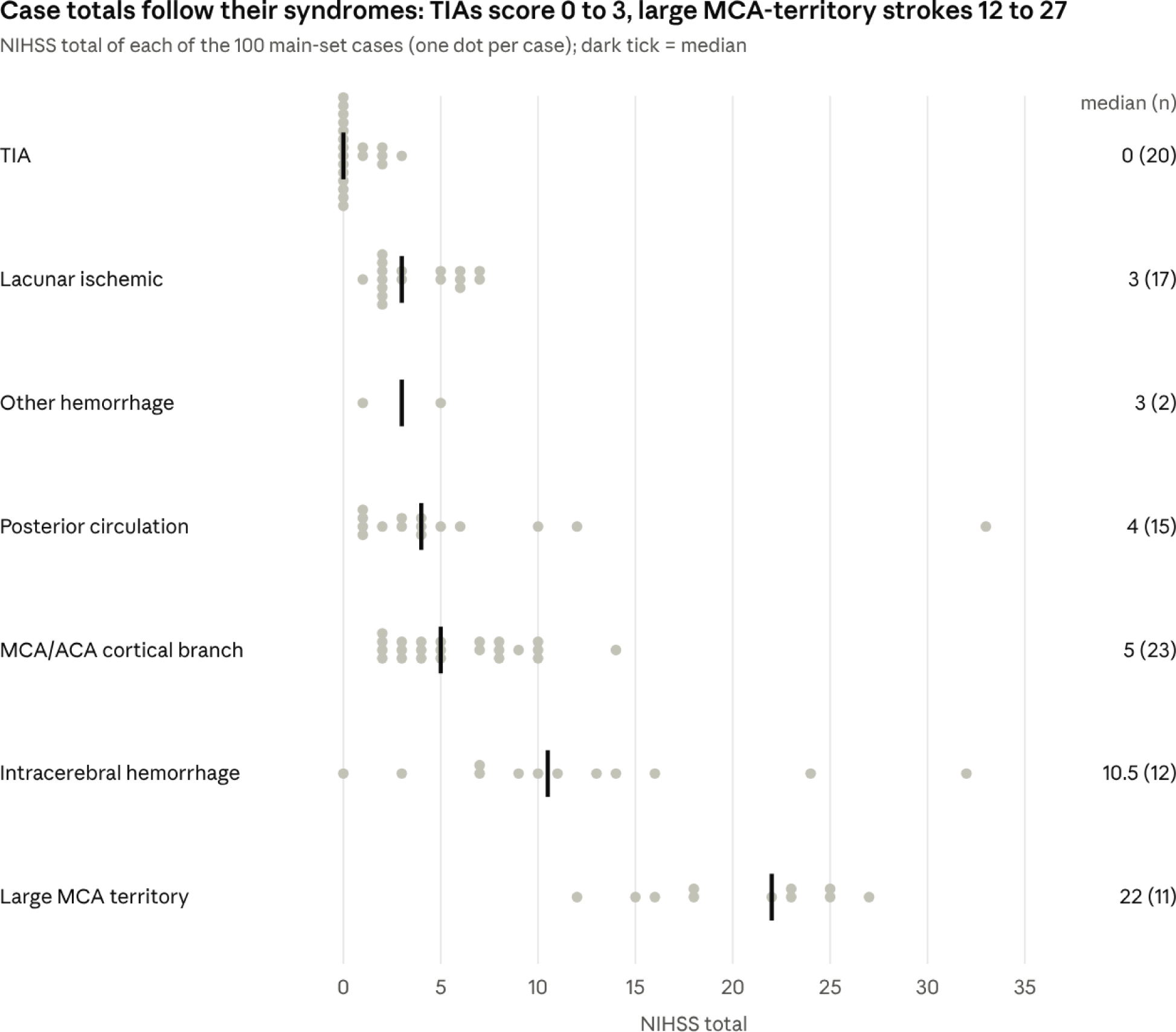
NIHSS totals by syndrome in the main set. Most TIAs scored 0 at the time of the exam (14 of 20), lacunar and posterior circulation strokes were mostly mild, and all large MCA-territory strokes scored 12 or more. Hemorrhage totals varied widely. The two high outliers outside the anterior circulation are a basilar occlusion (33) and a pontine hemorrhage (32).

**Figure 3.**
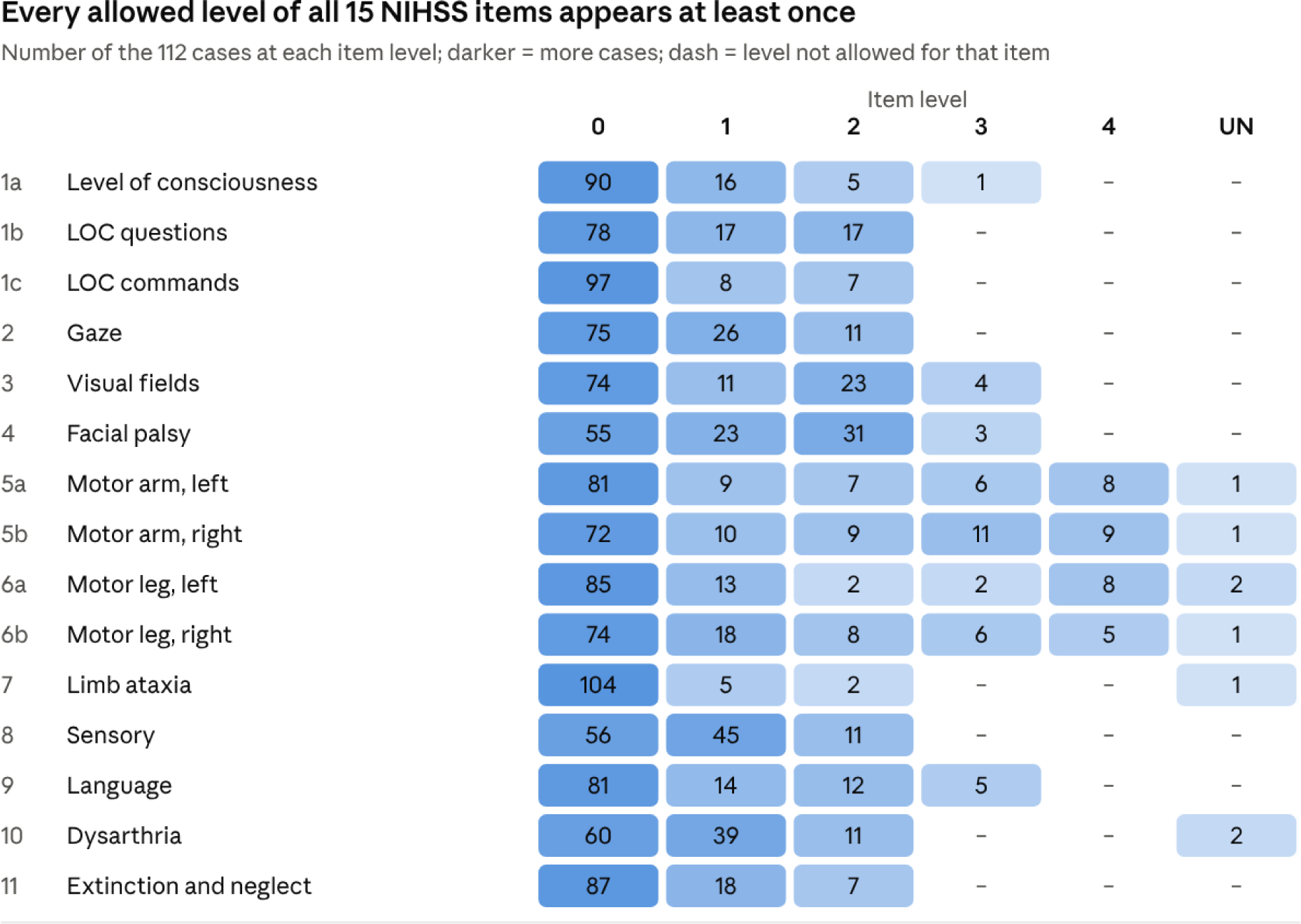
Coverage of NIHSS item levels across all 112 cases. Every level allowed for each item was scored at least once, including rare levels such as coma (1a = 3), bilateral blindness (3 = 3), untestable limbs and untestable speech. Most of the rarest levels came from the 12-case stress set.

#### Box 1. Opening of one transcript (case M021, narrated style)

Left MCA inferior-division (Wernicke-type) stroke in a 67-year-old woman. True scores: 1b = 2, 1c = 1, visual fields = 2, sensory = 1, language = 2, all other items 0; total 8.

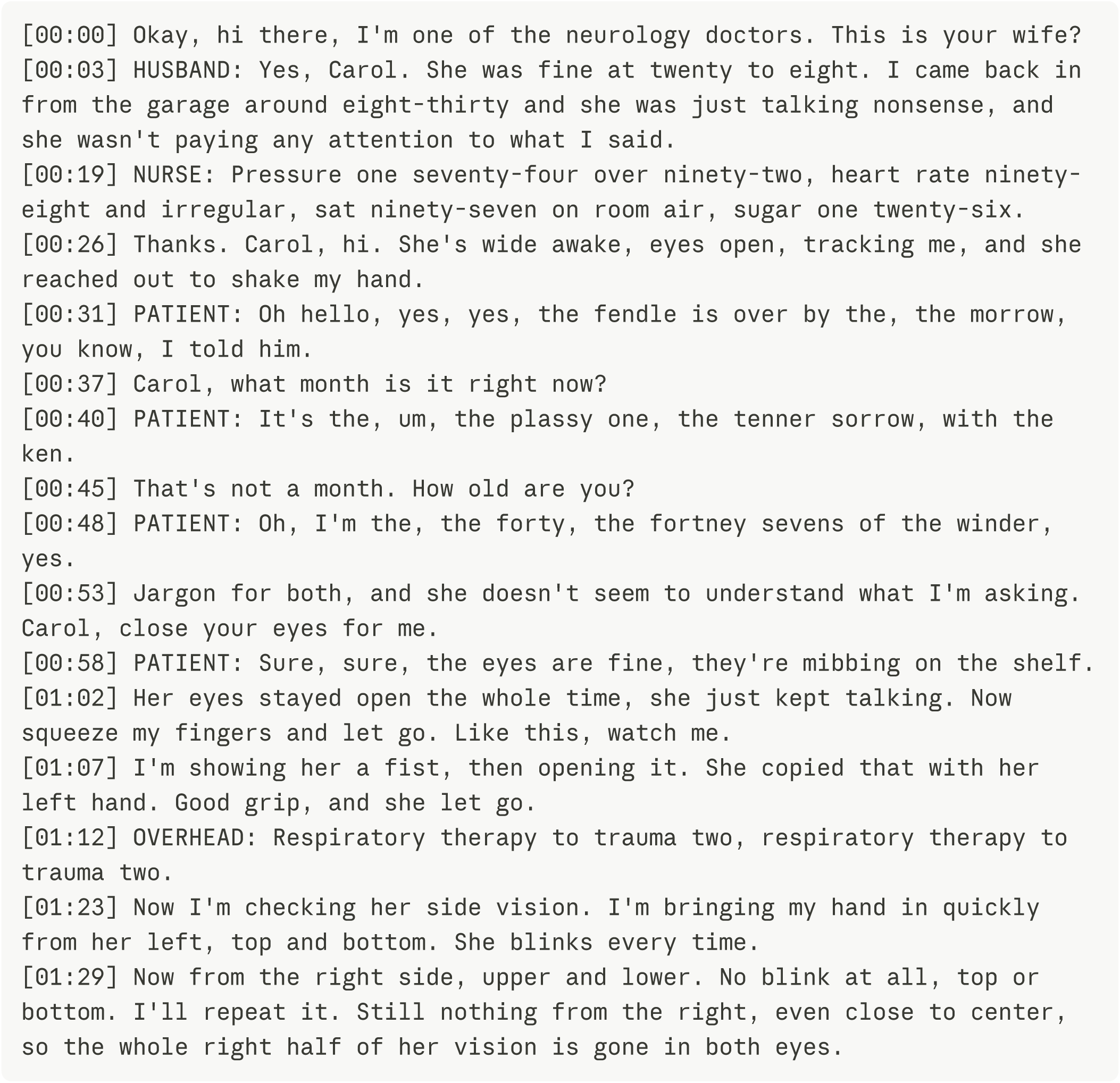

Each finding appears through what the patient says and does (jargon answers, a failed verbal command with an imitated grip, no blink to threat on the right) rather than as a score. Lines at 00:12, 00:14 and 01:16 are omitted from this excerpt. Across all 112 transcripts, a second model reading them blind recovered 99.2% of item scores (see Reference standard), which indicates that the findings can be recovered from the text alone.

### Primary outcomes

The LLM scored items more accurately than Jev (99.0% vs 94.4%; McNemar p < 0.001), whereas Jev was about 40 times faster at one third of the cost. Neither engine failed on any transcript.

**Table 3.** Accuracy, speed and cost on 112 clean transcripts.

| Measure | Jev | gpt-6-luna |
| --- | --- | --- |
| Item accuracy, % (95% CI) | 94.4 (93.2–95.4) | 99.0 (98.4–99.4) |
| Exact total score, n/112 | 63 | 101 |
| Total off by 1 point, n/112 (too high / too low) | 27 (21 / 6) | 10 (3 / 7) |
| Total off by 2 points, n/112 (too high / too low) | 18 (10 / 8) | 1 (0 / 1) |
| Total off by 3 points or more, n/112 | 4 (largest 7) | 0 |
| Total within 1 point, n/112 | 90 | 111 |
| Mean absolute total error, points | 0.72 | 0.11 |
| Largest total error, points | 7 | 2 |
| Latency, median / 95th percentile, s | 0.15 / 0.29 | 5.78 / 11.77 |
| Cost per 1,000 exams, USD | 0.23 | 0.67 |

On paired items, 10 were scored correctly only by Jev and 87 only by the LLM. At the case level, the LLM got more items right in 47 transcripts and Jev in 3, with 62 ties (Wilcoxon p < 0.001), and the accuracy difference was 4.6 percentage points (95% CI 3.2 to 6.1). Jev answered “not assessed” for 8 items that the transcript had in fact tested, which the LLM never did. When Jev’s total was wrong, it was more often too high than too low (33 vs 16 cases).

### Accuracy by item

**Figure 4.**
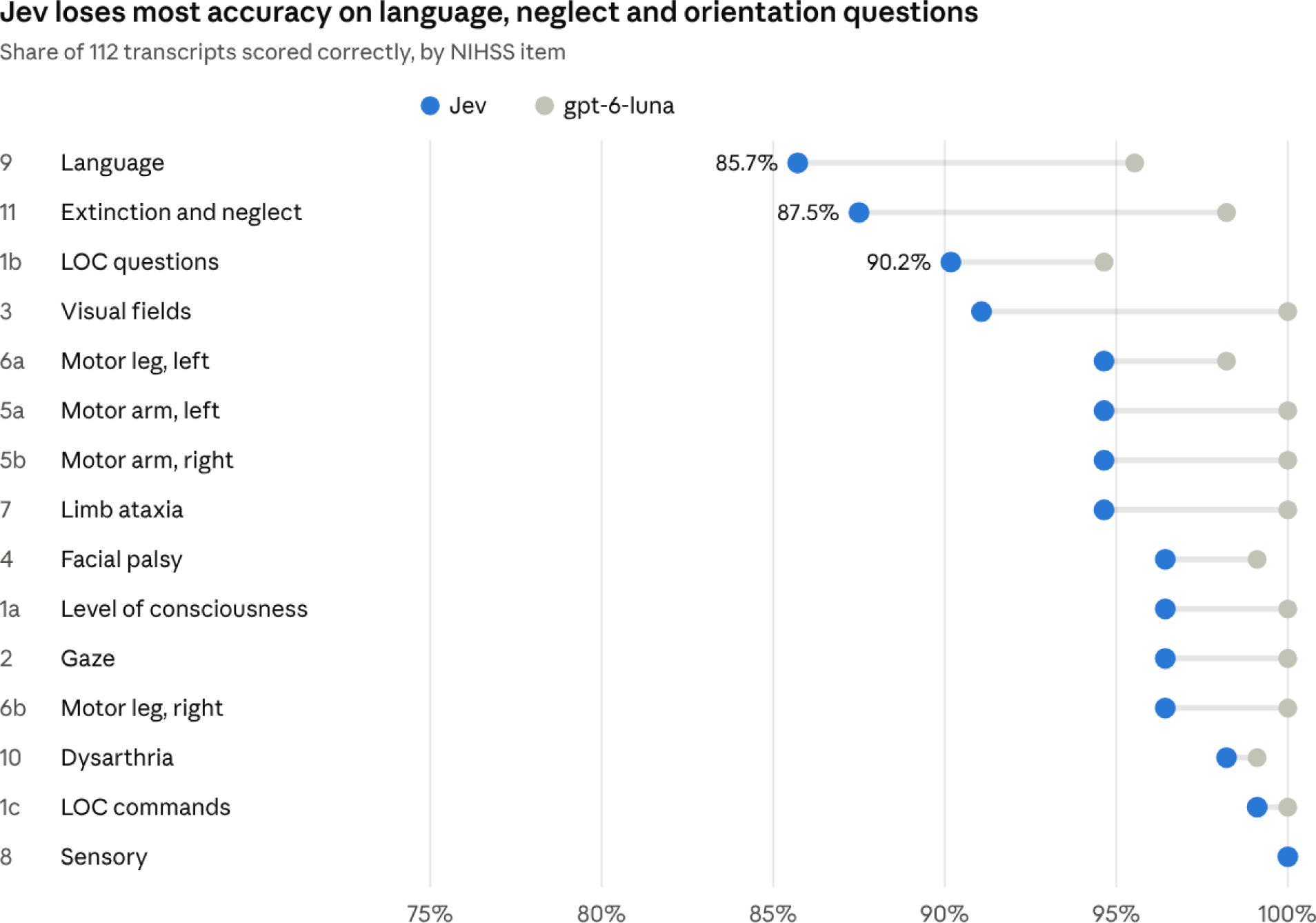
Jev’s accuracy was lowest for language (item 9, 85.7%), extinction and neglect (item 11, 87.5%), LOC questions (item 1b, 90.2%) and visual fields (item 3, 91.1%). The LLM reached at least 94.6% on every item, with its lowest accuracy on LOC questions (94.6%) and language (95.5%). Both engines scored sensory correctly in all 112 cases.

### Jev error patterns

On its four weakest items, Jev tended to repeat the same type of mistake (Table 4), and most of these errors occurred in telegraphic transcripts, where the examiner leaves the finding implied.

**Table 4.**
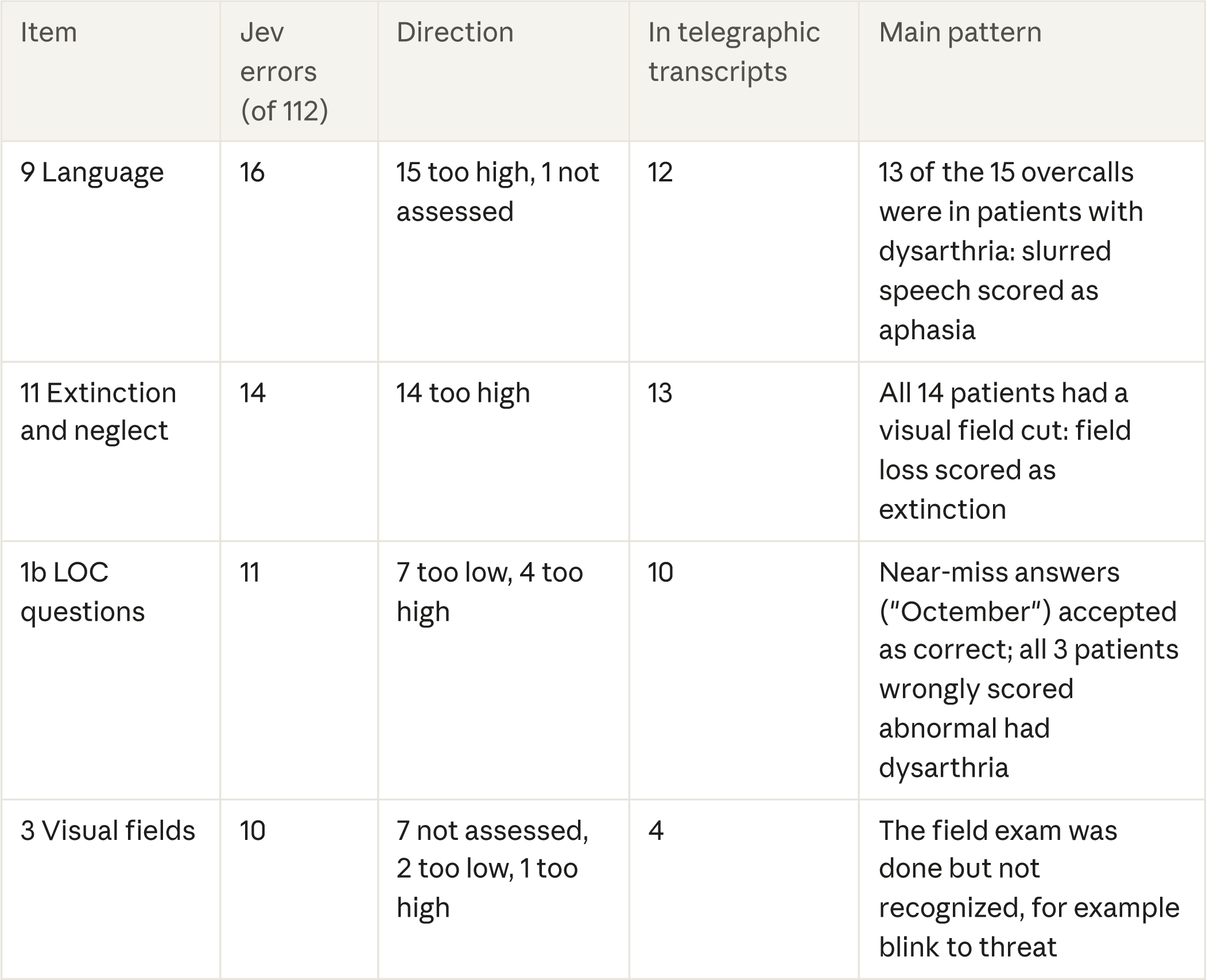
Jev error patterns on the four weakest items.

The LLM made far fewer errors on these items (5 on item 9, 2 on item 11, 6 on item 1b and none on item 3).

**Figure 5.**
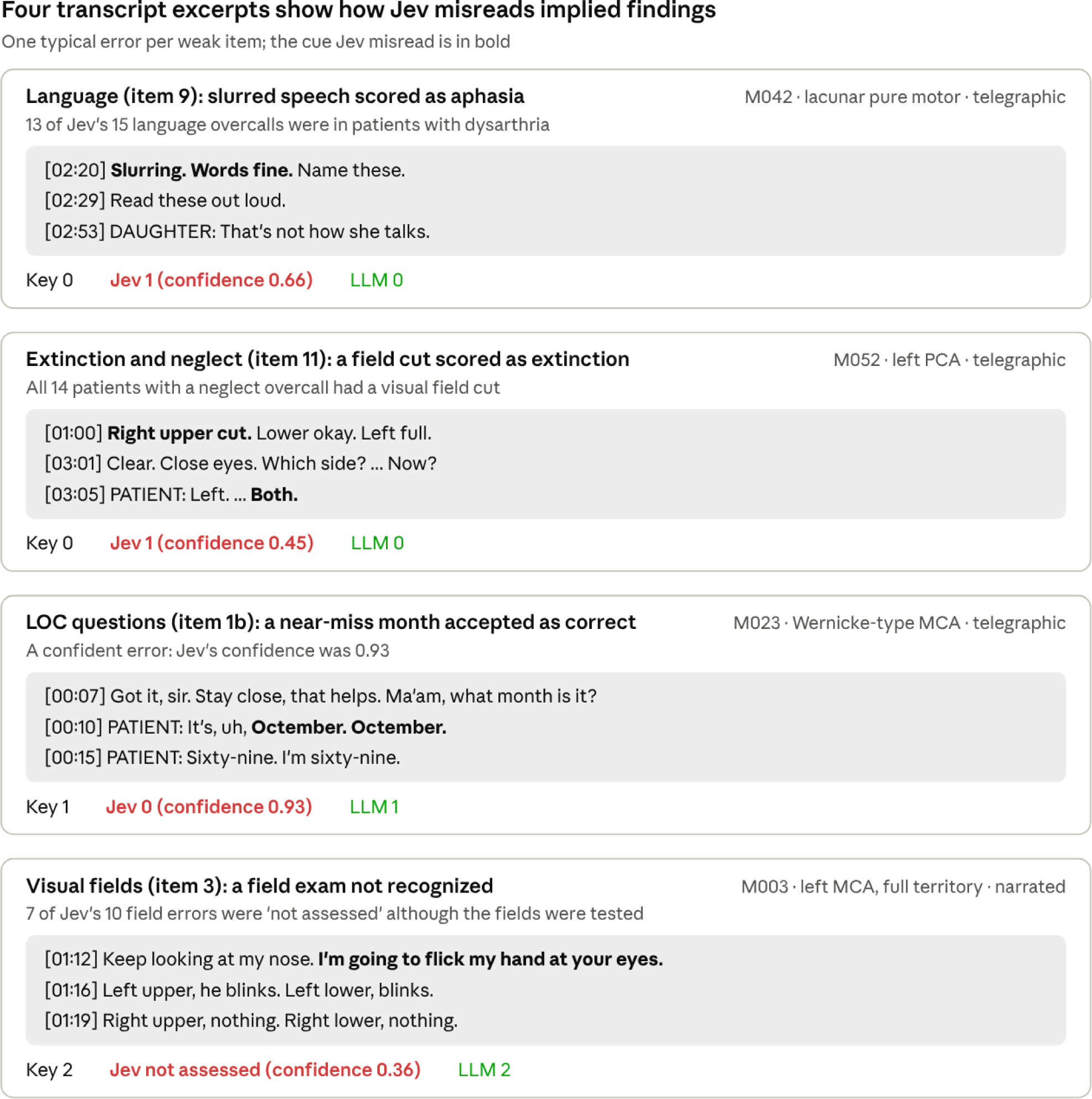
In each excerpt the finding is present in the transcript but stated briefly or shown indirectly. Jev tended to read a nearby abnormality, such as slurring or a field cut, as a deficit on the neighboring item, and to miss a test that was not named. Lines are quoted exactly, except that one line between the two questions is omitted in the M023 excerpt.

### Accuracy by examiner style

**Figure 6.**
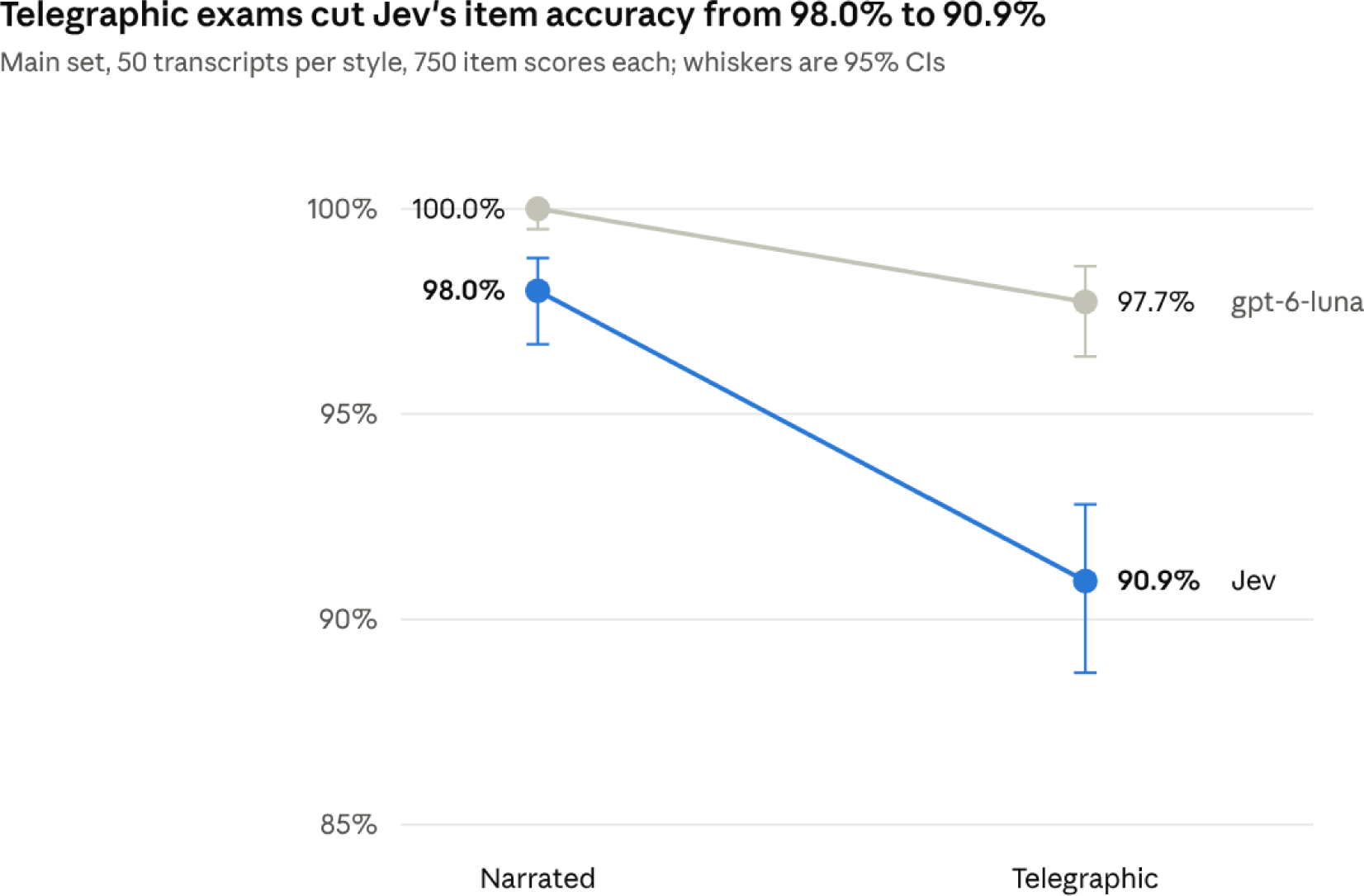
In the main set, where styles were split evenly, Jev’s item accuracy was 7.1 points lower when the examiner spoke in clipped shorthand, and its exact totals fell from 38/50 to 20/50. The LLM’s accuracy was 2.3 points lower (exact totals 50/50 vs 39/50). Stress cases were all narrated and are excluded from this comparison. In the stress set, the LLM scored all 180 items correctly and Jev 169 (93.9%).

### Jev confidence and a simulated hybrid (exploratory)

Jev’s confidence separated right from wrong answers, with a mean of 0.91 on correct items and 0.53 on incorrect ones. The LLM’s self-reported confidence separated them less well (0.97 vs 0.78).

**Figure 7.**
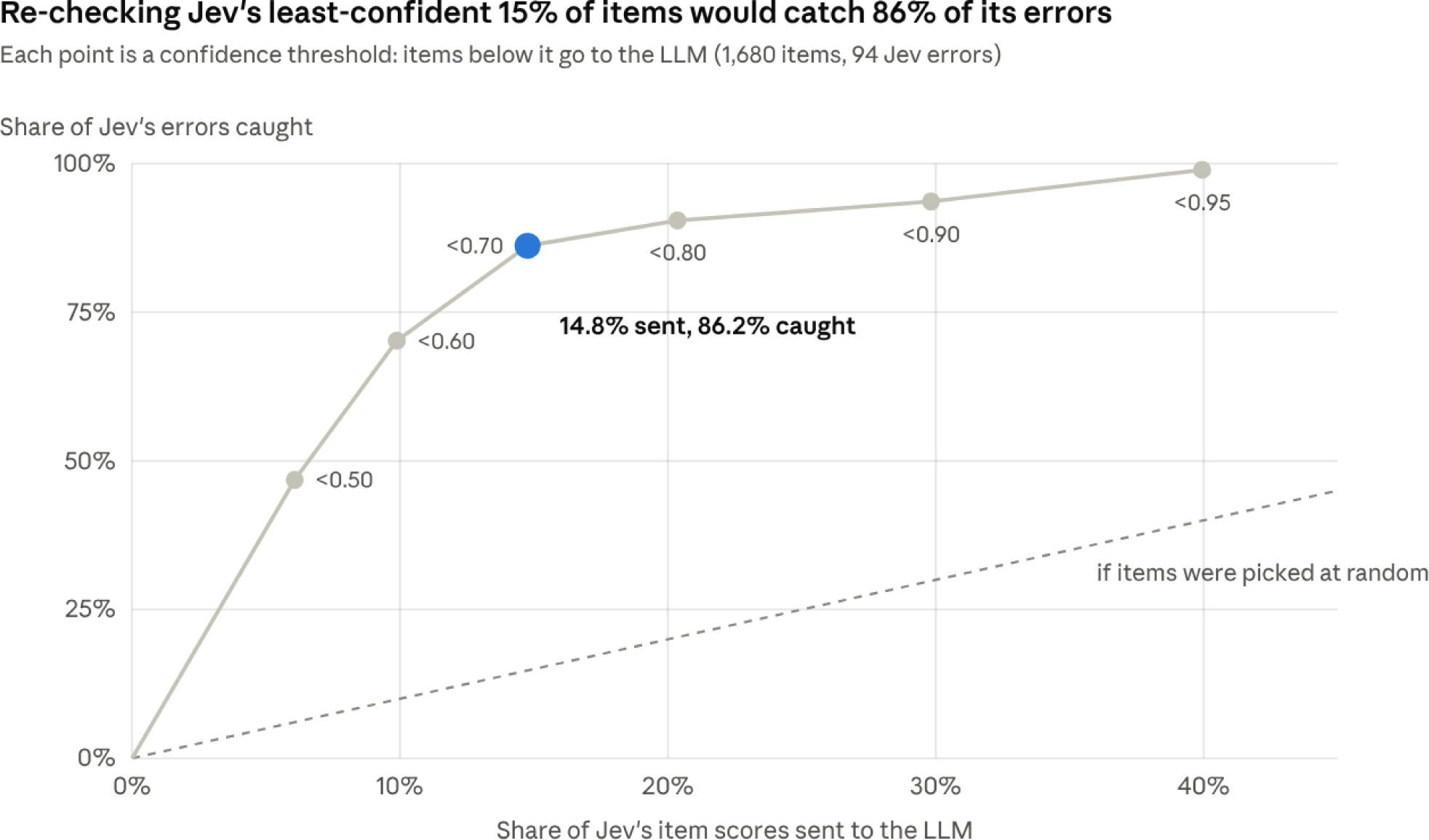
Sending to the LLM only the items on which Jev’s confidence was below 0.70 (14.8% of items) would have caught 81 of Jev’s 94 errors. In a post-hoc simulation using LLM answers already collected, this hybrid reached 98.8% item accuracy (1659/1680), close to the LLM alone (99.0%), while querying the LLM for about one item in seven. A live hybrid run, in which the LLM sees only the items passed to it, has not yet been done.

### Examiner-only transcripts

When only the examiner’s words were kept, item accuracy fell from 94.5% to 89.1% for Jev and from 99.0% to 90.4% for the LLM (Table 5). How much was lost depended on examiner style. With a narrating examiner, both engines stayed close to their full-transcript accuracy (Jev 96.9%, LLM 98.8%), whereas with a telegraphic examiner both fell to about 80%. The largest drops were on items whose evidence comes from the patient, above all the LOC questions on month and age (Jev 90.2% to 62.5%, LLM 94.6% to 60.7%), and sensory, language and extinction also fell. Items that the examiner observes and states, such as gaze, facial palsy, limb motor and ataxia, changed little.

**Table 5.** Item accuracy with full and examiner-only transcripts (jev-1.14.0 in both conditions).

| Examiner style | Jev, full | Jev, examiner-only | LLM, full | LLM, examiner-only |
| --- | --- | --- | --- | --- |
| Narrated (n = 62) | 97.3% | 96.9% | 100% | 98.8% |
| Telegraphic (n = 50) | 90.9% | 79.5% | 97.7% | 80.0% |
| All (n = 112) | 94.5% | 89.1% | 99.0% | 90.4% |

Without the patient’s speech, the LLM was no longer more accurate than Jev (73 items correct by Jev only vs 95 by the LLM only; McNemar χ² 2.63, p = 0.10). Exact totals fell to 58 of 112 for Jev and 74 of 112 for the LLM.

## Discussion

In this synthetic benchmark, a decision model and an LLM both scored most NIHSS items correctly from code-stroke exam transcripts. The LLM was more accurate at the item level and far more often exactly right on the total (101 vs 63 of 112). The decision model was about 40 times faster at about one third of the cost, and its confidence flagged most of its own errors.

Jev’s errors were concentrated on a few items and followed recognizable patterns. On its four weakest items, it tended to read a nearby finding as a deficit on the neighboring item. It scored slurred speech as aphasia and a visual field cut as extinction, accepted a near-miss answer (”Octember”) as correct, and marked a field test it did not recognize by name as “not assessed”. These are the items where NIHSS rules separate closely related findings. Because the errors are consistent, they may be correctable by changing how each question is put to the engine, without retraining it.

Examiner style mattered more for Jev than for the LLM. When the examiner stated findings aloud, Jev was nearly as accurate as the LLM (98.0% vs 100%), but with clipped shorthand and implied findings its accuracy fell to 90.9%. An ambient tool may therefore work best when examiners say their findings out loud, which may also help with teaching and handover. For dysarthria, an audio version would need either the examiner to name it or a separate sound-based detector.

The examiner-only analysis supports this. With a narrating examiner, the physician’s words alone cost either engine only about 1 percentage point of accuracy, which suggests that a single microphone on the examiner, avoiding much of the noise and crosstalk of the emergency department, may be sufficient. Such a setup would miss the patient’s own answers, which carry the LOC questions, language and sensory findings, unless the examiner repeats them aloud (”says March, incorrect”) or the patient’s voice is captured on a second channel.

The confidence analysis points to a practical design, in which a decision model gives an immediate score and passes only its uncertain items to an LLM. In our data, routing the 14.8% of items with confidence below 0.70 would have caught 81 of Jev’s 94 errors and, in a post-hoc simulation, raised accuracy to 98.8%, close to the LLM alone, with an LLM call for roughly one item in seven. This design still has to be tested in a live hybrid run.

Accuracy in our study was higher than in a study that extracted NIHSS scores from discharge summaries [5], but the tasks differ. Our transcripts were written to contain every finding, whereas clinical notes are often incomplete. Certified human raters reach item-level kappas of 0.15 to 0.90 on certification material [2,3]. Agreement with a fixed key and agreement between raters are different measures, however, so these numbers cannot be compared directly with ours.

### Limitations

- **Synthetic cases and text.** Cases, score forms and transcripts were all generated by an LLM. LLM-written text may be easier for an LLM to score than real bedside speech, which could favor the LLM engine.
- **Model-verified key.** The key was checked by a second model from the same company as the generator (Anthropic) and has not yet been reviewed by human raters. A prespecified blind review of 44 transcripts by two certified raters will be reported in a revised version.
- **No audio.** Transcripts were clean text, without real speech-recognition errors, crosstalk or noise.
- **Phonetic slurring.** Slurred words were spelled phonetically in the patient’s lines, which speech-to-text software would not preserve. With real audio, dysarthria, like the visual findings (gaze, fields, face, limb drift and ataxia), could be scored only if the examiner says it aloud. Some of Jev’s language errors, in which slurred speech was scored as aphasia, may partly reflect how the text was written.
- **Limited variation.** We tested two examiner styles, one LLM and one decision model, with a single run per engine and a fixed encounter month.
- **Engine versions.** TypeSafe serves only its current Jev release. The main results used jev-1.13.0, which can no longer be selected, so the run cannot be repeated exactly. The examiner-only analysis was post hoc.
- **Scope.** Stroke mimics were excluded, so the study tests scoring, not diagnosis.
- **Exploratory hybrid.** The hybrid analysis was post hoc and used LLM answers that had already been collected, not a live hybrid system.

### Conclusions

A decision model and an LLM can both score the NIHSS from exam transcripts with high item accuracy. The LLM was more accurate, while the decision model was faster and cheaper and could flag its own uncertainty, which supports a hybrid design. The next step is to test both on recorded, unscripted exams scored by human raters.

## Declarations

### Ethics

No human participants, patient data or identifiable information were used; all cases were synthetic. Institutional review board approval was therefore not required.

### Funding

None. Model and API costs were paid by WA.

### Competing interests

The authors declare no competing interests and have no financial relationship with TypeSafe (Jev), OpenAI or Anthropic beyond standard paid subscriptions and API use.

### Data and code availability

All score forms, transcripts, prompts, full model responses and analysis scripts are available from the corresponding author on request. To keep the planned human rater review blind, they will be made public at https://github.com/waleedabood/nihss-transcript-benchmark once that review is complete.

### Use of generative AI

Claude Opus 5.5 (Anthropic) generated the presentation table, score forms and transcripts. Claude Fable 5.1 (Anthropic) performed the blind checks of the key. Jev (TypeSafe) and gpt-6-luna (OpenAI) are the systems under test. Claude (Anthropic) assisted with analysis code, figures, drafting and language editing. The authors reviewed all content and take full responsibility for it.

### Author contributions

NA critically reviewed the manuscript and verified the statistical analysis. WA conceived and designed the study, made all design decisions, interpreted the results, and wrote the manuscript. Both authors approved the final manuscript.

## Data Availability

All score forms, transcripts, prompts, full model responses and analysis scripts are available from the corresponding author on reasonable request. To keep a planned blind human rater review unbiased, they will be made publicly available at https://github.com/waleedabood/nihss-transcript-benchmark once that review is complete.

## Notes

### Competing Interest Statement

The authors have declared no competing interest.

